## Supplementary Data for "Sodium channel-inhibiting drugs and cancer-specific survival: a population-based study of electronic primary care data"

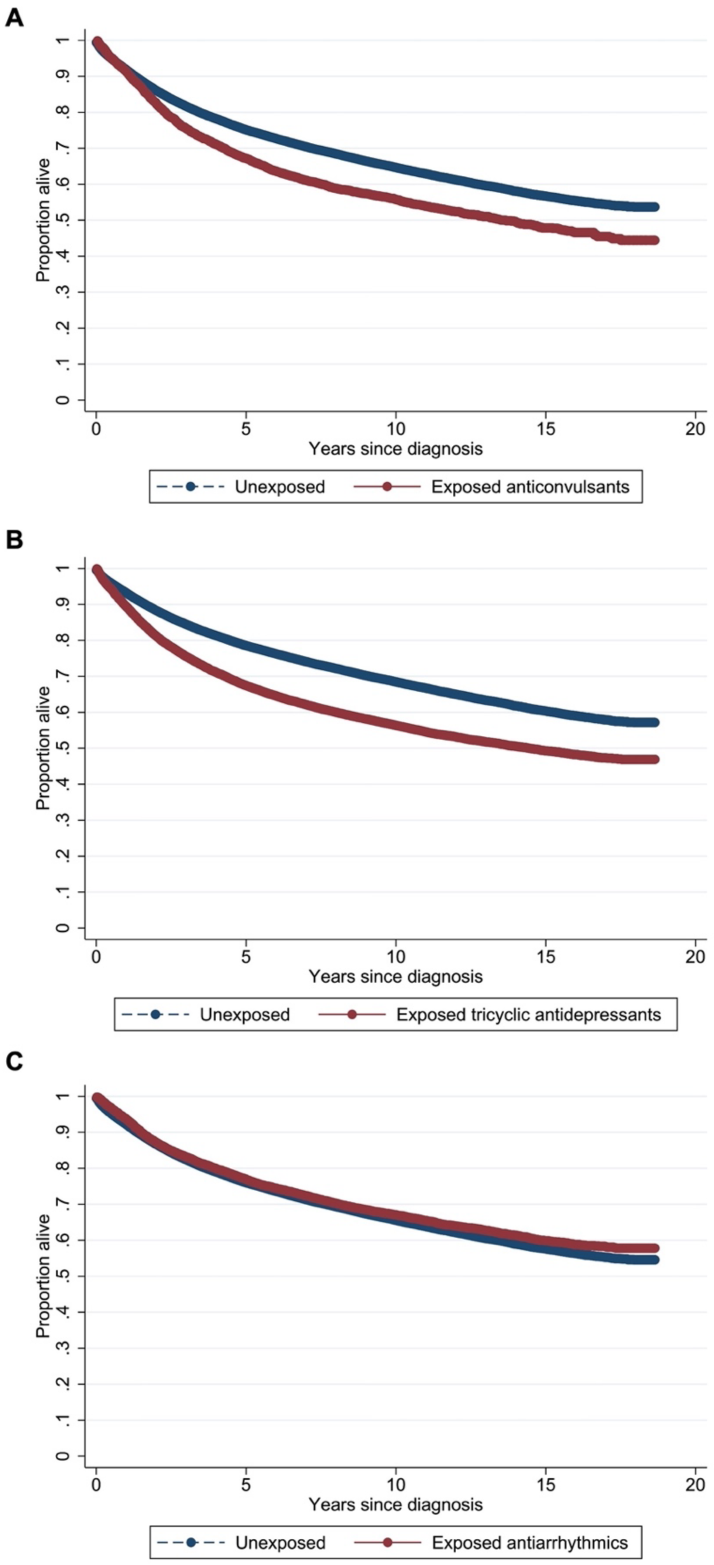

**Supplementary Figure 1.** Simon-Makuch survival curves for unexposed cancer patients and those ever exposed to VGSC-inhibiting anticonvulsant (A), tricyclic antidepressant (B) and antiarrhythmic (C) drugs in Scenario 3.

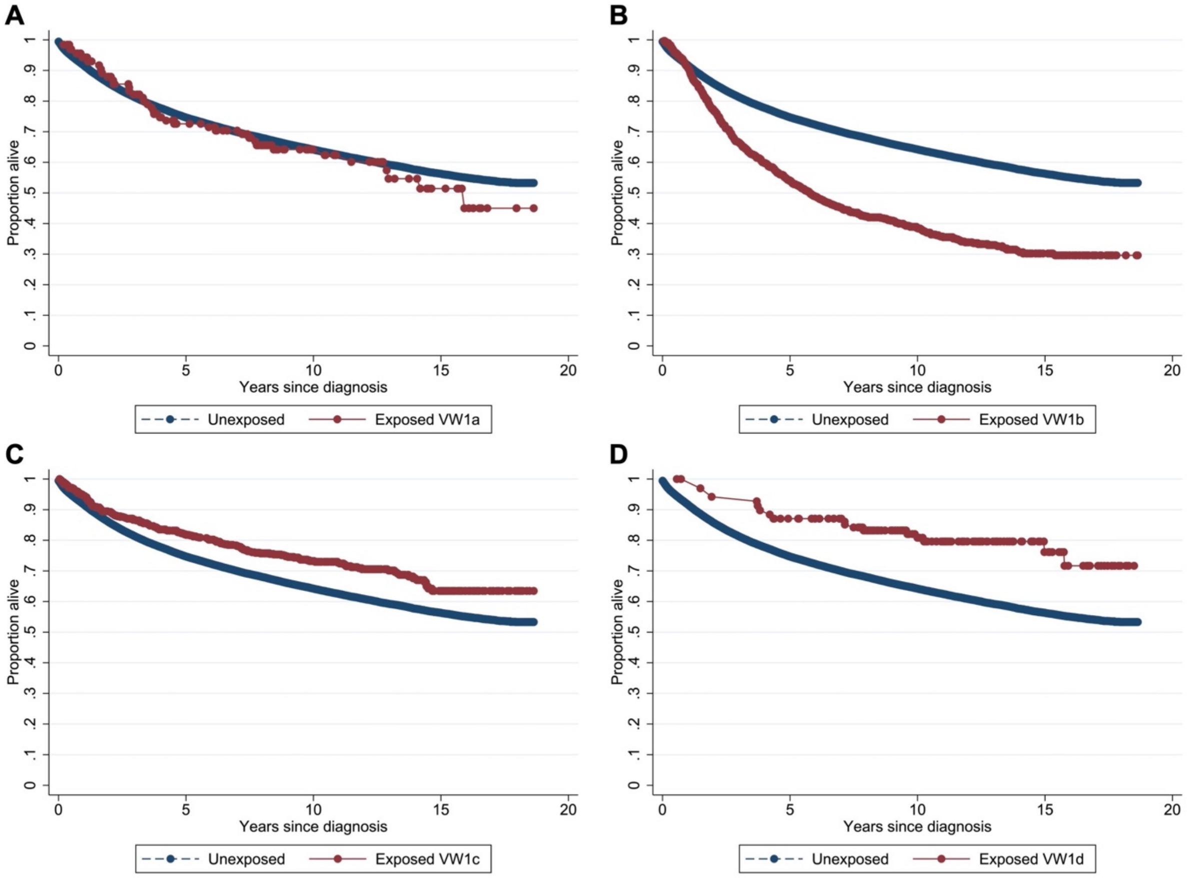

**Supplementary Figure 2.** Simon-Makuch survival curve for unexposed cancer patients and those ever exposed to Vaughan-Williams Class 1a (A), 1b (B), 1c (C) and 1d (D) drugs in Scenario 3.

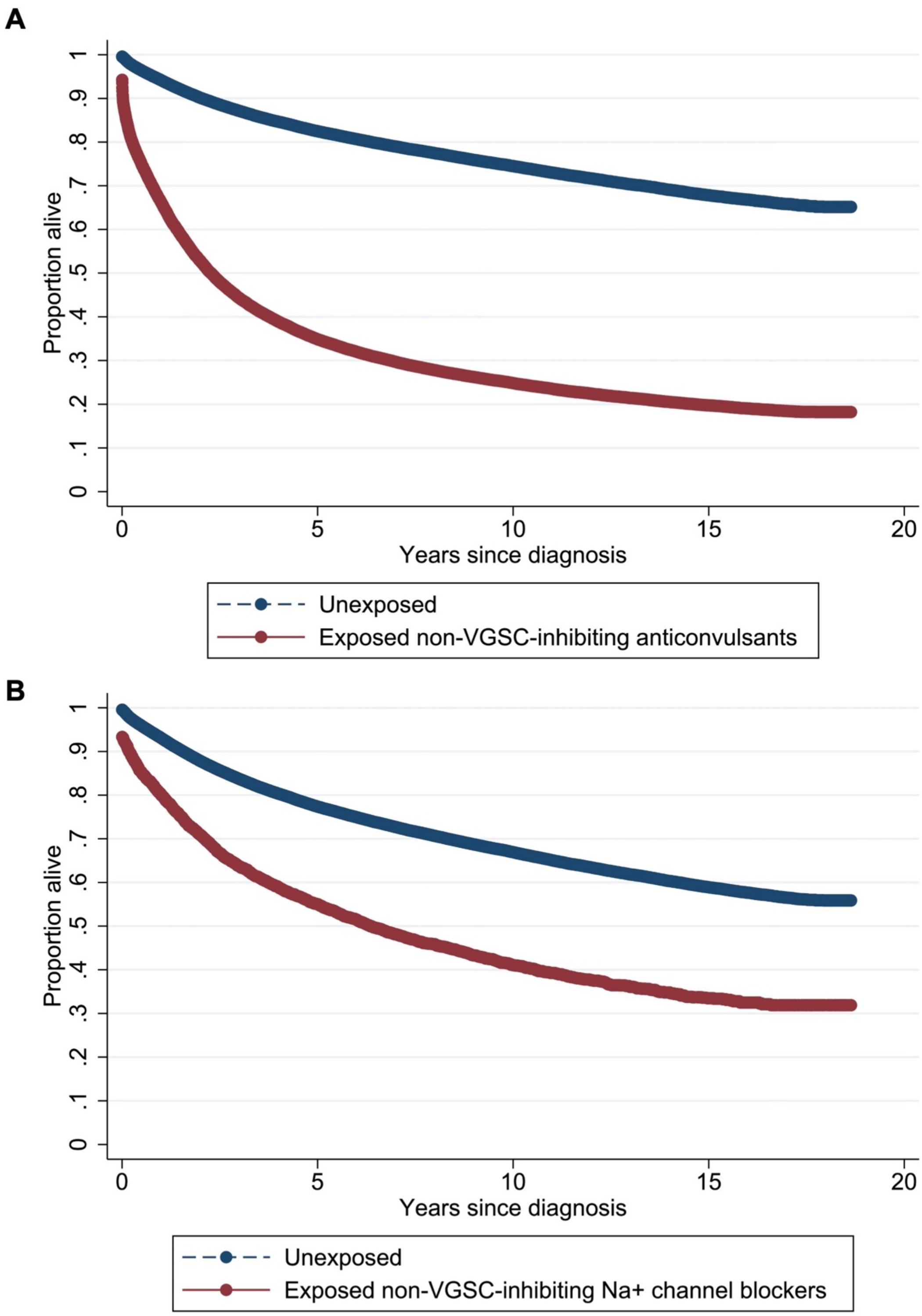

**Supplementary Figure 3.** Simon-Makuch survival curve for unexposed cancer patients and those ever exposed to non-VGSC-inhibiting anticonvulsants (A) and non-VGSC-inhibiting $Na^{+}$ channel blockers (B) in Scenario 3.

**Supplementary Table 1. Drug groups and classifications used in this study.**

| Drug | Classification | Vaughan Williams Classification[a] |
|---|---|---|
| **A. VGSC inhibitors** | | |
| Articaine | Amide local anaesthetic | |
| Bupivacaine | Amide local anaesthetic | |
| Cinchocaine | Amide local anaesthetic | |
| Etidocaine | Amide local anaesthetic | |
| Levobupivacaine | Amide local anaesthetic | |
| Lidocaine | Amide local anaesthetic | 1b |
| Mepivacaine | Amide local anaesthetic | |
| Prilocaine | Amide local anaesthetic | |
| Ropivacaine | Amide local anaesthetic | |
| Trimecaine | Amide local anaesthetic | |
| Ranolazine | Antiarrhythmic | 1d |
| Ajmaline | Antiarrhythmic | 1a |
| Amiodarone | Antiarrhythmic | 3 |
| Aprindine | Antiarrhythmic | 1b |
| Disopyramide | Antiarrhythmic | 1a |
| Dronedarone | Antiarrhythmic | 3 |
| Encainide | Antiarrhythmic | 1c |
| Flecainide | Antiarrhythmic | 1c |
| Mexiletine | Antiarrhythmic | 1b |
| Moricizine/Moricizine hydrochloride | Antiarrhythmic | 1c |
| Pilsicainide | Antiarrhythmic | 1c |
| Procainamide | Antiarrhythmic | 1a |
| Propafenone | Antiarrhythmic | 1c |
| Quinidine | Antiarrhythmic | 1a |
| Tocainide | Antiarrhythmic | 1b |

| | | |
|---|---|---|
| Carvedilol | Antiarrhythmic | 2 |
| Labetalol | Antiarrhythmic | 2 |
| Oxprenolol | Antiarrhythmic | 2 |
| Propranolol | Antiarrhythmic | 2 |
| Esmolol | Antiarrhythmic | 2 |
| Carbamazepine | Anticonvulsant | |
| Eslicarbazepine | Anticonvulsant | |
| Eslicarbazepine acetate | Anticonvulsant | |
| Ethotoin | Anticonvulsant | |
| Fosphenytoin | Anticonvulsant | |
| Lacosamide | Anticonvulsant | |
| Lamotrigine | Anticonvulsant | |
| Oxcarbazepine | Anticonvulsant | |
| Phenytoin | Anticonvulsant | 1b |
| Rufinamide | Anticonvulsant | |
| Sodium Valproate | Anticonvulsant | |
| Topiramate | Anticonvulsant | |
| Valproic acid | Anticonvulsant | |
| Zonisamide | Anticonvulsant | |
| Benzocaine | Ester local anaesthetic | |
| Procaine | Ester local anaesthetic | |
| Tetracaine | Ester local anaesthetic | |
| Riluzole | ALS treatment | |
| Amitriptyline | Tricyclic antidepressant | |
| Desipramine | Tricyclic antidepressant | |
| Duloxetine | Tricyclic antidepressant | |
| Fluoxetine | Tricyclic antidepressant | |
| Imipramine | Tricyclic antidepressant | |
| Maprotiline | Tricyclic antidepressant | |

| | |
|---|---|
| Nortriptyline | Tricyclic antidepressant |
| **A. Non-VGSC $Na^+$ channel blocker** | |
| Amiloride | ENaC inhibitor |
| Triamterene | ENaC inhibitor |
| **B. Non-VGSC-inhibiting anticonvulsants** | |
| Perampanel | AMPA receptor non-competitive antagonist |
| Stiripentol | Aromatic allylic alcohol |
| Phenobarbital | Barbiturate |
| Primidone | Barbiturate |
| Clobazam | Benzodiazepine |
| Clonazepam | Benzodiazepine |
| Diazepam | Benzodiazepine |
| Lorazepam | Benzodiazepine |
| Midazolam | Benzodiazepine |
| Ethosuximide | $Ca^{2+}$ channel inhibitor |
| Gabapentin | $Ca^{2+}$ channel inhibitor |
| Pregabalin | $Ca^{2+}$ channel inhibitor |
| Acetazolamide | Carbonic anhydrase inhibitor |
| Tiagabine | GABA reuptake inhibitor |
| Vigabatrin | GABA reuptake inhibitor |
| Brivaracetam | SV2A inhibitor |
| Levetiracetam | SV2A inhibitor |

[a]According to (37,38).

**Supplementary Table 2**. Characteristics of the participants stratified by exposure status.

| | **A VGSC-inhibitor prescription (any at any time exc anaesthetics) (n=53724)** | **No VGSC-inhibitor prescriptions (n=79272)** | **Total (n=132996)** | **p-value** |
|---|---|---|---|---|
| **Sex, n (%)** | | | | |
| Male | 22531 (41.9) | 41402 (52.2) | 63933 (48.1) | <0.001 |
| Female | 31193 (58.1) | 37870 (47.8) | 69063 (51.9) | |
| **Age at diagnosis, years** | | | | |
| Mean (SD) | 65.9 (13.0) | 68.0 (13.3) | 67.1 (13.2) | <0.001 |
| **Ethnicity, n (%)** | | | | |
| White | 50495 (94.0) | 72056 (90.9) | 122551 (92.1) | <0.001 |
| Mixed/Multiple ethnic groups | 161 (0.3) | 299 (0.4) | 460 (0.3) | |
| Asian/Asian British | 801 (1.5) | 1297 (1.6) | 2098 (1.6) | |
| Black/Black British | 733 (1.4) | 1597 (2.0) | 2330 (1.8) | |
| Other | 190 (0.4) | 336 (0.4) | 526 (0.4) | |

| | | | | |
|---|---|---|---|---|
| Not recorded/known | 1344 (2.5) | 3687 (4.7) | 5031 (3.8) | |
| **Index of Multiple Deprivation, n (%)** | | | | |
| Mean (SD) | 9.2 (5.6) | 9.0 (5.5) | 9.1 (5.5) | <0.001 |
| **Smoking status, n (%)** | | | | |
| Heavy smoker | 1596 (3.0) | 1648 (2.1) | 3244 (2.4) | <0.001 |
| Moderate smoker | 4503 (8.4) | 5259 (6.6) | 9762 (7.3) | |
| Light smoker | 1675 (3.1) | 2156 (2.7) | 3831 (2.9) | |
| Ex-smoker | 16413 (30.6) | 23004 (29.0) | 39417 (29.6) | |
| Non-smoker | 28676 (53.4) | 44072 (55.6) | 72748 (54.7) | |
| Not recorded/known | 861 (1.6) | 3133 (4.0) | 3994 (3.0) | |
| **Alcohol intake, n (%)** | | | | |
| Heavy drinker | 13656 (25.4) | 20565 (25.9) | 34221 (25.7) | <0.001 |
| Moderate drinker | 3421 (6.4) | 5003 (6.3) | 8424 (6.3) | |
| Light drinker | 11829 (22.0) | 16231 (20.5) | 28060 (21.1) | |

| | | | | |
|---|---|---|---|---|
| Non drinker | 9001 (16.8) | 10442 (13.2) | 19443 (14.6) | |
| Not recorded/known | 15817 (29.4) | 27031 (34.1) | 42848 (32.2) | |
| **BMI category, n (%)** | | | | |
| Overweight/Obese | 32166 (59.9) | 42432 (53.5) | 74598 (56.1) | <0.001 |
| Normal range | 15526 (28.9) | 24137 (30.4) | 39663 (29.8) | |
| Underweight | 2345 (4.4) | 3598 (4.5) | 5943 (4.5) | |
| Not recorded/known | 3687 (6.9) | 9105 (11.5) | 12792 (9.6) | |
| **Physical activity, n (%)** | | | | |
| Very active | 2634 (4.9) | 4173 (5.3) | 6807 (5.1) | <0.001 |
| Moderately active | 18614 (34.6) | 25806 (32.6) | 44420 (33.4) | |
| Inactive | 7551 (14.1) | 8179 (10.3) | 15730 (11.8) | |
| Not recorded/known | 24925 (46.4) | 41114 (51.9) | 66039 (49.7) | |
| **Type of cancer, n (%)** | | | | |
| Breast | 27106 (50.5) | 32422 (40.9) | 59528 (44.8) | <0.001 |

| | | | | |
|---|---|---|---|---|
| Bowel | 8435 (15.7) | 14432 (18.2) | 22867 (17.2) | |
| Prostate | 18183 (33.8) | 32418 (40.9) | 50601 (38.0) | |
| **Total CCI score** | | | | |
| Mean (SD) | 6.1 (2.8) | 5.9 (2.7) | 6.0 (2.7) | <0.001 |
| **VGSC-inhibitor indication[a], n (%)** | | | | |
| Epilepsy | 1915 (3.6) | 449 (0.6) | 2364 (1.8) | <0.001 |
| Cardiac arrhythmia | 9646 (18.0) | 9791 (12.4) | 19437 (14.6) | <0.001 |
| Amyotrophic lateral sclerosis | 0 (0.0) | 0 (0.0) | 0 (0.0) | - |
| Neuropathic pain/painful neuropathy | 9860 (18.4) | 7271 (9.2) | 17131 (12.9) | <0.001 |
| **≥1 of above, n (%)** | 18744 (34.9) | 16048 (20.2) | 34792 (26.2) | <0.001 |

[a] not mutually exclusive

VGSC, voltage gated sodium channel; SD, standard deviation; BMI, body mass index; CCI, Charlson Comorbidity Index score.

**Supplementary Table 3**. Characteristics of the 'ever' exposed group stratified by timing of exposure relative to their cancer diagnosis.

| | **Before only** (n=14,157) | **Before and after** (n=17,264) | **After only** (n=22,303) | **VGSC inhibitor prescription (any at any time excluding local anaesthetics)** (n=53,724) |
|---|---|---|---|---|
| **Sex, n (%)** | | | | |
| Male | 6049 (42.7) | 6325 (36.6) | 10157 (45.5) | 22531 (41.9) |
| Female | 8108 (57.3) | 10939 (63.4) | 12146 (54.5) | 31193 (58.1) |
| **Age at diagnosis, years** | | | | |
| Mean (SD) | 68.6 (13.4) | 65.7 (12.9) | 64.3 (12.5) | 65.9 (13.0) |
| **Ethnicity, n (%)** | | | | |
| White | 13189 (93.2) | 16366 (94.8) | 20940 (93.9) | 50495 (94.0) |
| Mixed/Multiple ethnic groups | 35 (0.2) | 47 (0.3) | 79 (0.4) | 161 (0.3) |
| Asian/Asian British | 172 (1.2) | 252 (1.5) | 377 (1.7) | 801 (1.5) |
| Black/Black British | 169 (1.2) | 186 (1.1) | 378 (1.7) | 733 (1.4) |
| Other | 48 (0.3) | 54 (0.3) | 88 (0.4) | 190 (0.4) |
| Not recorded/known | 544 (3.8) | 359 (2.1) | 441 (2.0) | 1344 (2.5) |

| | | | | |
|---|---|---|---|---|
| **Index of Multiple Deprivation, n (%)** | | | | |
| Mean (SD) | 9.2 (5.5) | 9.5 (5.7) | 9.0 (5.5) | 9.2 (5.6) |
| **Smoking status, n (%)** | | | | |
| Heavy smoker | 373 (2.6) | 561 (3.2) | 662 (3.0) | 1596 (3.0) |
| Moderate smoker | 1139 (8.0) | 1554 (9.0) | 1810 (8.1) | 4503 (8.4) |
| Light smoker | 437 (3.1) | 520 (3.0) | 718 (3.2) | 1675 (3.1) |
| Ex-smoker | 4336 (30.6) | 5362 (31.1) | 6715 (30.1) | 16413 (30.6) |
| Non-smoker | 7562 (53.4) | 9020 (52.2) | 12094 (54.2) | 28676 (53.4) |
| Not recorded/known | 310 (2.2) | 247 (1.4) | 304 (1.4) | 861 (1.6) |
| **Alcohol intake, n (%)** | | | | |
| Heavy drinker | 3467 (24.5) | 4030 (23.3) | 6159 (27.6) | 13656 (25.4) |
| Moderate drinker | 929 (6.6) | 1036 (6.0) | 1456 (6.5) | 3421 (6.4) |
| Light drinker | 3153 (22.3) | 3879 (22.5) | 4797 (21.5) | 11829 (22.0) |
| Non drinker | 2436 (17.2) | 3404 (19.7) | 3161 (14.2) | 9001 (16.8) |
| Not recorded/known | 4172 (29.5) | 4915 (28.5) | 6730 (30.2) | 15817 (29.4) |
| **BMI category, n (%)** | | | | |

| | | | | |
|---|---|---|---|---|
| Overweight/Obese | 7990 (56.4) | 10699 (62.0) | 13477 (60.4) | 32166 (59.9) |
| Normal range | 4267 (30.1) | 4734 (27.4) | 6525 (29.3) | 15526 (28.9) |
| Underweight | 764 (5.4) | 728 (4.2) | 853 (3.8) | 2345 (4.4) |
| Not recorded/known | 1136 (8.0) | 1103 (6.4) | 1448 (6.5) | 3687 (6.9) |
| **Physical activity, n (%)** | | | | |
| Very active | 615 (4.3) | 752 (4.4) | 1267 (5.7) | 2634 (4.9) |
| Moderately active | 4497 (31.8) | 5896 (34.2) | 8221 (36.9) | 18614 (34.6) |
| Inactive | 1785 (12.6) | 2816 (16.3) | 2950 (13.2) | 7551 (14.1) |
| Not recorded/known | 7260 (51.3) | 7800 (45.2) | 9865 (44.2) | 24925 (46.4) |
| **Type of cancer, n (%)** | | | | |
| Breast | 6766 (47.8) | 9642 (55.9) | 10698 (48.0) | 27106 (50.5) |
| Bowel | 2531 (17.9) | 2472 (14.3) | 3432 (15.4) | 8435 (15.7) |
| Prostate | 4860 (34.3) | 5150 (29.8) | 8173 (36.6) | 18183 (33.8) |
| **Total CCI score** | | | | |
| Mean (SD) | 6.3 (2.8) | 6.1 (2.8) | 5.9 (2.8) | 6.1 (2.8) |
| **Diagnosis of a VGSC inhibitor indication[a], n (%)** | | | | |

| | | | | |
|---|---|---|---|---|
| Epilepsy | 202 (1.4) | 1271 (7.4) | 442 (2.0) | 1915 (3.6) |
| Cardiac arrhythmia | 2611 (18.4) | 3362 (19.5) | 3673 (16.5) | 9646 (18.0) |
| Amyotrophic lateral sclerosis | 0 (0.0) | 0 (0.0) | 0 (0.0) | 0 (0.0) |
| Neuropathic pain/painful neuropathy | 2126 (15.0) | 3790 (22.0) | 3944 (17.7) | 9860 (18.4) |
| **≥1 of above, n (%)** | 4400 (31.1) | 7199 (41.7) | 7145 (32.0) | 18744 (34.9) |
| **Died, n (%)** | 7800 (55.1) | 8119 (47.0) | 10109 (45.3) | 26028 (48.4) |
| **Most common prescription 1[a]** | | | | |
| Tricyclic antidepressant | 9312 (65.8) | 11495 (66.6) | 17793 (79.8) | 38600 (71.8) |
| Antiarrhythmic | 4023 (28.4) | 3828 (22.2) | 3181 (14.3) | 11032 (20.5) |
| Anticonvulsant | 822 (5.8) | 1935 (11.2) | 1305 (5.9) | 4062 (7.6) |
| Treatment for ALS | 0 (0.0) | 6 (0.0) | 24 (0.1) | 30 (0.1) |
| **Most common prescription 2[b]** | | | | |
| Tricyclic antidepressant | 7887 (55.7) | 10831 (62.7) | 15187 (68.1) | 33905 (63.1) |
| Antiarrhythmic | 3604 (25.5) | 3726 (21.6) | 2881 (12.9) | 10211 (19.0) |
| Amide local anaesthetic | 1905 (13.5) | 782 (4.5) | 2951 (13.2) | 5638 (10.5) |

| | | | | |
|---|---|---|---|---|
| Anticonvulsant | 708 (5.0) | 1907 (11.0) | 1191 (5.3) | 3806 (7.1) |
| Ester local anaesthetic | 53 (0.4) | 12 (0.1) | 71 (0.3) | 136 (0.3) |
| Treatment for ALS | 0 (0.0) | 6 (0.0) | 22 (0.1) | 28 (0.1) |
| **Length of exposure (days)** | | | | |
| Mean (SD) | 2095.1 (2220.1) | 4889.9 (2473.2) | 1912.7 (2077.5) | 2917.5 (2627.5) |
| < 6 months, n (%) | 3239 (22.9) | 40 (0.2) | 4589 (20.6) | 7868 (14.6) |
| ≥ 6 months, n (%) | 10918 (77.1) | 17224 (99.8) | 17714 (79.4) | 45856 (85.4) |
| **Recent exposure[c], n (%)** | 1017 (7.2) | 8580 (49.7) | 0 (0.0) | 9597 (17.9) |

[a] excluding local anaesthetics.

[b] including local anaesthetics.

[c] ≥2 prescriptions relating to one of the VGSC-inhibiting drugs within 2 years before the date of the cancer diagnosis, including at least one within 6 months before.

**Supplementary Table 4.** Deaths stratified by exposure to non-VGSC-inhibiting anticonvulsants and non-VGSC-inhibiting $Na^+$ channel blockers.

| | **A VGSC-inhibitor prescription (any other than Amide or Ester local anaesthetics at any time) (n=53724)** | **No VGSC-inhibitor prescriptions (except Amide or Ester local anaesthetics) (n=79272)** | **A non-VGSC-inhibiting anticonvulsant prescription (any at any time) (n=46017)** | **No exposure to a non-VGSC-inhibiting anticonvulsant prescription (n=86979)** | **Non-VGSC-inhibiting $Na^+$ channel blocker prescription (any at any time) (n=9256)** | **No exposure to a non-VGSC-inhibiting $Na^+$ channel blocker prescription (n=123740)** |
|---|---|---|---|---|---|---|
| Died (any cause) | 26028 (48.4) | 40932 (51.6) | 25284 (54.9) | 41676 (47.9) | 6969 (75.3) | 59991 (48.5) |
| Died with any cancer as underlying cause | 15933 (29.7) | 26104 (32.9) | 17987 (39.1) | 24050 (27.7) | 3601 (38.9) | 38436 (31.1) |
| Died with any cancer as contributory cause | 18598 (34.6) | 30492 (38.5) | 20003 (43.5) | 29087 (33.4) | 4540 (49.0) | 44550 (36.0) |
| Died with index cancer as underlying cause | 12282 (22.9) | 20443 (25.8) | 13925 (30.3) | 18800 (21.6) | 2842 (30.7) | 29883 (24.1) |

| | | | | | | |
|---|---|---|---|---|---|---|
| Died with index cancer as contributory cause | 15256 (28.4) | 25482 (32.1) | 16451 (35.7) | 24287 (27.9) | 3834 (41.4) | 36904 (29.8) |

**Supplementary Table 5.** Estimates of the relationship between exposure to non-VGSC-inhibiting drugs, subdivided by type, and cancer-specific mortality (empty cells indicate that analysis was not permitted due to low numbers).

| **Non-VGSC-inhibiting anticonvulsants drug groups** | **Exposed* (n=46017), n (%)** | **HR (95% CI) p-value Scenario 1** | **HR (95% CI) p-value Scenario 2** | **HR (95% CI) p-value Scenario 3** |
|---|---|---|---|---|
| *Ever use* | | | | |
| AMPA receptor non-competitive antagonist | 4 (0.1) | - | - | - |
| Aromatic allylic alcohol | 0 (0.0) | - | - | - |
| Barbiturate | 469 (1.0) | 1.39 (1.19, 1.63) p<0.001 | 1.28 (1.07, 1.52) p=0.01 | 1.42 (1.19, 1.69) p<0.001 |
| Benzodiazepine | 37696 (81.9) | 3.01 (2.95, 3.07) p<0.001 | 3.73 (3.65, 3.81) p<0.001 | 4.91 (4.80, 5.02) p<0.001 |
| Calcium channel inhibitor | 11643 (21.7) | 2.19 (2.12, 2.26) p<0.001 | 2.27 (2.19, 2.34) p<0.001 | 2.80 (2.71, 2.90) p<0.001 |
| Carbonic anhydrase inhibitor | 14274 (31.0) | 0.92 (0.79, 1.06) p=0.25 | 0.93 (0.76, 1.13) p=0.46 | 1.25 (1.02, 1.53) p=0.03 |
| GABA reuptake inhibitor | 23 (0.1) | - | - | - |
| SV2A inhibitor | 620 (1.4) | 2.98 (2.63, 3.38) p<0.001 | 3.03 (2.67, 3.44) p<0.001 | 3.75 (3.30, 4.25) p<0.001 |
| *Recent use* | | | | |

| | | | | |
|---|---|---|---|---|
| AMPA receptor non-competitive antagonist | 0 (0.0) | - | - | - |
| Aromatic allylic alcohol | 0 (0.0) | - | - | - |
| Barbiturate | 208 (0.5) | 1.44 (1.16, 1.78) p<0.001 | 1.37 (1.10, 1.71) p<0.001 | 1.46 (1.18, 1.82) p<0.001 |
| Benzodiazepine | 2686 (5.8) | 1.46 (1.37, 1.56) p<0.001 | 1.34 (1.25, 1.44) p<0.001 | 1.51 (1.41, 1.62) p<0.001 |
| Calcium channel inhibitor | 902 (2.0) | 1.12 (0.99, 1.26) p=0.08 | 1.06 (0.93, 1.21) p=0.38 | 1.20 (1.05, 1.37) p=0.01 |
| Carbonic anhydrase inhibitor | 43 (0.1) | - | - | - |
| GABA reuptake inhibitor | 0 (0.0) | - | - | - |
| SV2A inhibitor | 54 (0.1) | - | - | - |
| *Most common VGSC-inhibitor prescription* | | | | |
| AMPA receptor non-competitive antagonist | 0 (0.0) | - | - | - |
| Aromatic allylic alcohol | 0 (0.0) | - | - | - |
| Barbiturate | 348 (0.8) | 1.43 (1.20, 1.71) p<0.001 | 1.30 (1.07, 1.59) p=0.01 | 1.40 (1.15, 1.71) p<0.001 |

| | | | | |
|---|---|---|---|---|
| Benzodiazepine | 32563 (70.8) | 3.06 (2.99, 3.12) p<0.001 | 4.02 (3.93, 4.11) p<0.001 | 5.11 (4.99, 5.23) p<0.001 |
| Calcium channel inhibitor | 12061 (26.2) | 2.22 (2.15, 2.30) p<0.001 | 2.33 (2.24, 2.41) p<0.001 | 2.80 (2.70, 2.90) p<0.001 |
| Carbonic anhydrase inhibitor | 561 (1.2) | 0.92 (0.78, 1.08) p=0.31 | 0.92 (0.72, 1.18) p=0.52 | 1.22 (0.96, 1.56) p=0.11 |
| GABA reuptake inhibitor | 7 (0.0) | - | - | - |
| SV2A inhibitor | 477 (1.0) | 3.16 (2.73, 3.65) p<0.001 | 3.23 (2.79, 3.74) p<0.001 | 4.00 (3.45, 4.63) p<0.001 |

*Figures in this column relate to the number of patients recorded as having at least some follow-up time considered as exposed to the drug group of interest in Scenario 1 for each definition (ever use, recent use, most common), as a percentage of the whole 'ever' exposed group. The number of patients with any person-time of follow-up considered as exposed for each drug group will be lower in Scenario 2, and fewer still in Scenario 3.
